# Coexisting autoantibodies against transcription factor Sp4 are associated with decreased cancer risk in dermatomyositis patients with anti-TIF1γ autoantibodies

**DOI:** 10.1101/2022.02.28.22271555

**Authors:** Yuji Hosono, Brandon Sie, Iago Pinal-Fernandez, Katherine Pak, Christopher A. Mecoli, Maria Casal-Dominguez, Blake M. Warner, Mariana J. Kaplan, Jemima Albayda, Sonye K. Danoff, Thomas E. Lloyd, Julie Paik, Eleni Tiniakou, Jose C. Milisenda, Josep M. Grau-Junyent, Albert Selva-O’Callaghan, Lisa Christopher-Stine, H. Benjamin Larman, Andrew L. Mammen

**Author notes:** **Address correspondence to:** Andrew L. Mammen, M.D., Ph.D., Muscle Disease Unit, Laboratory of Muscle Stem Cells and Gene Expression, National Institute of Arthritis and Musculoskeletal and Skin Diseases, National Institutes of Health, 50 South Drive, Room 1141, Building 50, MSC 8024, Bethesda, MD 20892. These authors contributed equally to this project (Yuji Hosono, Brandon Sie, Iago Pinal-Fernandez, Katherine Pak, H. Benjamin Larman, Andrew L. Mammen).

## Abstract

**Objectives:** In dermatomyositis (DM), different autoantibodies are associated with unique clinical phenotypes. For example, anti-TIF1γ autoantibodies are associated with a substantially increased risk of cancer. The purpose of this study was to discover novel DM autoantibodies.

**Methods:** Phage ImmunoPrecipitation Sequencing using sera from 67 DM patients suggested that transcription factor Sp4 is a novel autoantigen; this was confirmed by showing that patient sera immunoprecipitated full-length Sp4 protein. Sera from 371 Johns Hopkins myositis patients (255 with DM, 28 with antisynthetase syndrome [ASyS], 40 with immune-mediated necrotizing myopathy [IMNM], 29 with inclusion body myositis [IBM], and 19 with polymyositis [PM]), 75 rheumatologic disease controls (25 with Sjogren’s syndrome, 25 with systemic lupus erythematosus, and 25 with rheumatoid arthritis), and 200 healthy comparators were screened for anti-SP4 autoantibodies by an enzyme-linked immune absorption assay. Serum from 23 Spanish TIF1γ-positive DM patients was also screened for anti-Sp4 autoantibodies

**Results:** Anti-Sp4 autoantibodies were present in 11.4% of DM and 8% of rheumatoid arthritis patients but not in any other clinical group. Among DM patients, 90% of anti-Sp4 autoantibodies were detected in patients with anti-TIF1γ autoantibodies. Among anti-TIF1γ-positive DM patients from Johns Hopkins and Spain, those with coexisting anti-Sp4 autoantibodies had a decreased risk of cancer (0% vs. 31%; p=0.001, Chi-squared test).

**Conclusions:** Anti-Sp4 autoantibodies are enriched in anti-TIF1γ-positive DM patients without cancer, suggesting that the development of an anti-Sp4 immune response may correlate with a relatively low risk of cancer in these patients.

**KEY MESSAGES:** *What is already known about this subject?:*

- Dermatomyositis patients with anti-TIF1γ autoantibodies have an increased risk of cancer.

*What does this study add?:*

- Anti-Sp4 autoantibodies are enriched in dermatomyositis patients with anti-TIF1γ autoantibodies.
- Anti-Sp4 autoantibodies are only found in dermatomyositis patients without cancer.
- Muscle strength is greater in dermatomyositis patients with anti-Sp4 autoantibodies.
- Anti-Sp4 autoantibodies are not present in patients with antisynthetase syndrome, immune-mediated necrotizing myopathy, or inclusion body myositis.
- Autoantibodies against Sp4 are absent in those with systemic lupus erythematosus or Sjogren’s syndrome and present in 8% of those with rheumatoid arthritis.

*How might this impact on clinical practice?:* Testing for anti-Sp4 autoantibodies may define a population of anti-TIF1γ-positive dermatomyositis patients without a substantially increased risk of cancer.

## INTRODUCTION

The idiopathic inflammatory myopathies (IIM) are a heterogeneous family of diseases that includes dermatomyositis (DM), immune-mediated necrotizing myopathy (IMNM), the antisynthetase syndrome (ASyS), and inclusion body myositis (IBM)^1^. Most patients with IIM have a myositis-specific autoantibody (MSA). Among those with DM, approximately 70% have an MSA recognizing either TIF1γ, NXP2, Mi2, or MDA5. Importantly, each MSA is associated with a unique clinical phenotype. For instance, DM patients with anti-TIF1γ autoantibodies have a substantially increased risk of cancer^2^ whereas those with anti-Mi2 autoantibodies do not^3^. Although MSAs are usually mutually exclusive, there are exceptions. For example, some anti-MDA5-positive DM patients develop a second MSA recognizing splicing factor proline/glutamine-rich (SFPQ); these patients have a decreased risk of arthritis compared to anti-MDA5-positive patients without anti-SFPQ autoantibodies^4^.

Here we utilized Phage ImmunoPrecipitation Sequencing (PhIP-Seq)^5, 6^ to identify novel autoantibodies in DM patients. This approach revealed that autoantibodies recognizing transcription factor Sp4 are a novel MSA in DM patients with co-existing anti-TIF1γ autoantibodies. Furthermore, we show that anti-Sp4 autoantibodies are associated with the subset of TIF1γ-positive DM patients who do not have cancer. These findings raise the possibility that an immune response directed at Sp4 may correlate with a greatly reduced risk of cancer in this patient population.

## PATIENTS AND METHODS

### Patients and serum samples

The <u>discovery cohort</u> consisted of 67 patients enrolled in the Johns Hopkins Myositis Center Longitudinal study between 2002 and 2016 with a diagnosis of DM based on the criteria of Bohan and Peter^7, 8^ whose serum tested negative for all MSAs by line blot (EUROLINE myositis profile).

The <u>screening cohort</u> included myositis patients enrolled in the Johns Hopkins Myositis Center Longitudinal Cohort study between 2002 and 2018. This included patients with DM based on the criteria of Bohan and Peter^7, 8^, ASyS defined by the presence of anti-Jo1 autoantibodies, IMNM defined by the presence of anti-SRP or anti-HMGCR autoantibodies, IBM defined by the Lloyd and Greenberg criteria, as well as PM patients defined as those who fulfilled the criteria of Bohan and Peter for PM but who did not have ASyS or IMNM. Patients were considered positive for autoantibodies recognizing Mi2, NXP2, TIF1γ, MDA5, Jo1, SRP, HMGCR, SAE, or PmScl if they tested positive by at least two immunologic techniques from among the following: ELISA (including the MBL anti-TIF1γ ELISA, RG-7854R), *in vitro* transcription and translation immunoprecipitation, line blotting (EUROLINE myositis profile), Phip-seq,^9^ immunoprecipitation from S35-labeled HeLa cell lysates, or immunoprecipitation blotting.^10-12^.

A <u>validation cohort</u> included serum from 23 DM patients from Spain who tested positive for anti-TIF1γ autoantibodies by both an in-house line blot as well as the EUROLINE myositis profile line blot.

Serum from 25 patients with rheumatoid arthritis, 25 patients with systemic lupus erythematosus, 25 patients with Sjögren’s syndrome, and 200 healthy controls, all enrolled in studies at the National Institutes of Health, were also screened by ELISA for anti-Sp4 autoantibodies. Additional healthy control samples used for the analysis of peptidome data were from subjects self-reported to be free of autoimmune disease.

In the screening cohort, muscle strength was evaluated by the examining physician using the Medical Research Council scale. This scale was transformed to Kendall’s 0-10 scale for analysis purposes as previously described.^13^ Serial strength measurements for each patient were made by the same physician but more than 10 different physicians contributed to the measurements. For the purposes of analyses, right- and left-side measurements for arm abduction and hip flexion strength were combined and the average was used for calculations (possible range 0–10). Serum creatine kinase (CK) levels were included for the longitudinal analysis if obtained within a period of 6 weeks before or after strength testing. Skin manifestations (i.e., heliotrope rash or Gottron’s sign), weakness, symptoms of esophageal involvement, antisynthetase syndrome-associated clinical features (e.g. mechanics hands, Raynaud’s phenomenon, arthritis, fever), and other clinical features were documented both retrospectively at the onset of the disease (by asking patients about features present at the onset of disease) and prospectively at each visit. Interstitial lung disease was defined through a multidisciplinary approach as recommended by the American Thoracic Society.^14^ Cancer-associated myositis was defined as a malignancy occurring within 3 years either before or after the onset of myositis symptoms.

### Standard protocol approvals and patient consents

This study was approved by the Institutional Review Boards at Johns Hopkins, the National Institutes of Health, and the Vall d’Hebron and Clinic Hospitals; written informed consent was obtained from each participant.

### Screening for autoantibodies by PhIP-Seq

The standard PhIP-Seq procedure^9^ was used to profile sera from 67 DM patients that were seronegative for Mi2, NXP2, TIF1γ or MDA5 by EUROLINE line blot (discovery cohort). Briefly, the IgG concentration of each serum sample was measured via ELISA assay, which allowed normalization of the IgG input (2 *μ*g per reaction) into the PhIP-Seq assay. Serum antibody was mixed with the 90-aa human peptidome library^15^ and incubated overnight. Antibody and antibody-bound phage were captured using protein A and protein G coated Dynal magnetic beads (Invitrogen #10002D & #10004D). The immunoprecipitated phage DNA library was then amplified using PCR, with sample-specific DNA barcodes added during a second PCR reaction. Pooled amplicons were sequenced using an Illumina NextSeq instrument. An informatics pipeline was used for sample demultiplexing and alignment. Data normalization was performed by comparison of each sample against a set of mock immunoprecipitations as described previously^16^. Finally, the normalized PhIP-Seq profiles were compared against a previously established PhIP-Seq database of healthy volunteers using a custom case-control analysis script to identify reactivities specifically associated with DM (https://brandonsie.github.io/phipcc/).

### Immunoprecipitation using ^35^S-labeled *in vitro* transcription/translated (IVTT) Sp4

DNA encoding full-length human Sp4 was purchased (Origene) and used in IVTT reactions (Promega), generating ^35^S-labeled Sp4 protein. Immunoprecipitation of radiolabeled Sp4 was performed using patient serum or a mouse monoclonal anti-Sp4 positive control antibody (sc-515738, Santa Cruz Biotechnology), as previously described^17^.Immunoprecipitates were reduced, boiled, subjected to electrophoresis on 10% sodium dodecyl sulfated-polyacrylamide gels, and visualized by fluorography.

### Anti-Sp4 ELISA

ELISA plates (96-well) were coated overnight at 4°C with 100 ng of human recombinant Sp4 protein (H00006671-P01, Abnova Corporation, Taipei, Taiwan) diluted in 100 *μ*L of phosphate-buffered saline (PBS). After washing the plates with PBS including 0.05% Tween-20 (PBS-T) and blocking with 300 µl of 5% bovine serum albumin (BSA) in PBS-T for 1 hour at 37°C, the plates were washed with PBS-T. 100 µl of diluted human serum samples (1:400 with 1% BSA/PBS-T) was added to each well and incubated for 1 hour at 37°C. After washing with PBS-T, 100 µl of HRP-labeled goat anti-human antibody (1:10,000, catalog# 109-036-088, Jackson ImmunoResearch Lab, PA, USA) was added and incubated for 30 minutes at 37°C in the dark. After washing the plate with PBS-T and PBS, 100 µl of SureBlue TMB microwell peroxidase enzyme substrate kit (95059-286, KPL, MA, USA) was added. Reactions were stopped after 8 minutes. The absorbance at 450 nm was determined. Test sample absorbances were normalized to the sera of an arbitrary positive control sample, a reference sample included in every ELISA. The cutoff for a normal anti-Sp4 autoantibody titer (0.29 arbitrary units was defined as the mean plus two standard deviations of the normalized absorbances of the 200 healthy comparators. This cutoff was determined to be optimal based on a graphical analysis of the normalized absorbances.

### Statistical analysis

Dichotomous variables were expressed as percentages and absolute frequencies, and continuous features were reported as means and standard deviations (SD). Pairwise comparisons for categorical variables between groups were made using the chi-square test or Fisher’s exact test, as appropriate. Student’s t-test was used to compare continuous variables among groups and paired t-test was used to compare the level of weakness of different muscle groups. CK, a highly positively skewed variable, was expressed as median, first, and third quartile for descriptive purposes, and was transformed through a base-10 logarithm for regression analysis.

Statistical analyses related to clinical variables were performed using Stata/MP 14.1. A 2-sided p-value of 0.05 or less was considered statistically significant with no adjustment for multiple comparisons.

### Patient and Public Involvement statement

Patients and the public were not involved in the design, conduct, reporting, or dissemination plans of the present research.

### Data availability statement

All data relevant to the study are either included in the article or will be shared upon request.

## RESULTS

### Identification of Sp4 as a novel autoantigen

In an effort to discover novel DM-associated autoantibodies, we assembled a cohort of sera from DM patients who were negative for MSAs by EUROIMMUN line blot, which tests for DM-specific autoantibodies including anti-TIF1γ, anti-NXP2, anti-Mi2, anti-MDA5, and anti-SAE. These 67 serum samples were used in the PhIP-Seq assay with a T7 phage display library spanning all open reading frames in the human genome as overlapping 90 amino acid peptides^15^. While PhIP-Seq does not detect antibodies directed against conformational or post-translational epitopes, the approach is unbiased and provides a higher resolution map of autoantibody binding specificities^18^. Among the 67 serum samples analyzed, 13 recognized from 1 to 4 peptides corresponding to Transcription factor Sp4 (Table 1). Recognition of non-overlapping epitopes is indicative of a polyclonal antibody response that is more likely to be antigen-driven.

**Table 1.**
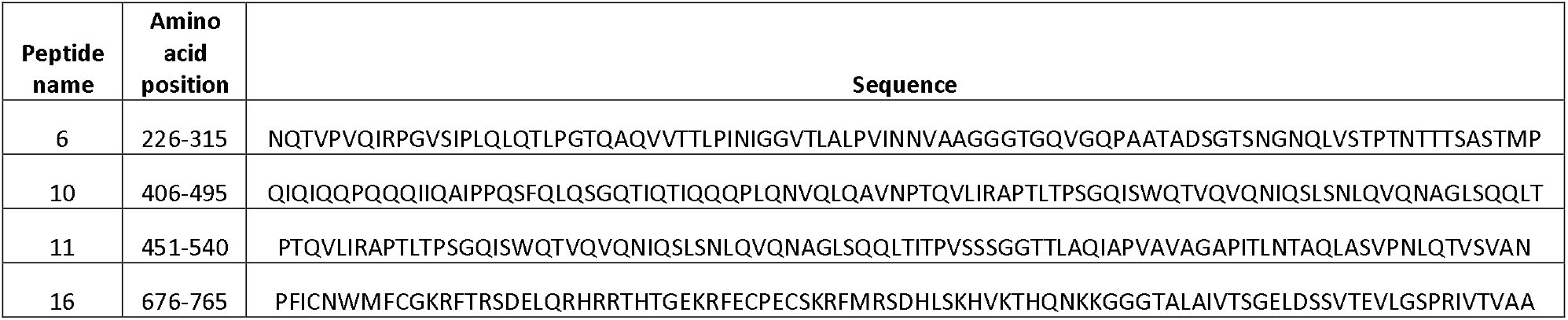
Sp4 peptides identified by PhIP-Seq.

To determine which sera had immunoreactivity against native Sp4, we generated full-length radiolabeled Sp4 protein by IVTT and used human serum or a rabbit anti-Sp4 positive control antibody to immunoprecipitate the protein. The positive control anti-Sp4 antibody and each of the 6 serum samples that recognized 3 or 4 distinct Sp4 peptides by PhIP-Seq (lanes 3-8) efficiently immunoprecipitated full-length Sp4 protein. In contrast, serum samples that recognized just 1 or 2 Sp4 peptides only weakly immunoprecipitated full-length Sp4 protein (lanes 9-15).

Unexpectedly, since they had each tested negative for anti-TIF1γ autoantibodies by EUROIMMUN line blot, we noted that all 6 serum samples that efficiently immunoprecipitated full-length Sp4 protein also recognized a peptide corresponding to TIF1γ by the PhIP-Seq assay (data not shown). This demonstrates the high sensitivity of PhIP-Seq and suggests that the line blot test may not be a sufficiently sensitive assay for detecting anti-TIF1γ autoantibodies. Indeed, a recent report showed that the EUROIMMUN line blot has good specificity but poor sensitivity for detecting anti-TIFγ autoantibodies compared to immunoprecipitation detection methods^19^.

### Screening for anti-Sp4 autoantibodies in patients with IIM, other rheumatologic conditions, and healthy controls

To screen patients rapidly for anti-Sp4 autoantibodies, we developed an ELISA. We defined a serum sample as being positive for anti-Sp4 autoantibodies if the relative absorbance was two standard deviations or higher than the mean value of 200 healthy control subjects. Using this method, we found that 6 of the 13 samples recognizing Sp4 peptides by PhIP-Seq were also ELISA positive; these were the same 6 samples that recognized 3 or 4 Sp4 peptides and which most efficiently immunoprecipitated full-length Sp4 protein (Figure 1, lanes 3-8). In contrast, among the 54 samples that did not recognize Sp4 peptides by PhIP-Seq, 50 were available for further testing and each of these was negative for anti-Sp4 autoantibodies by ELISA.

**Figure 1.**
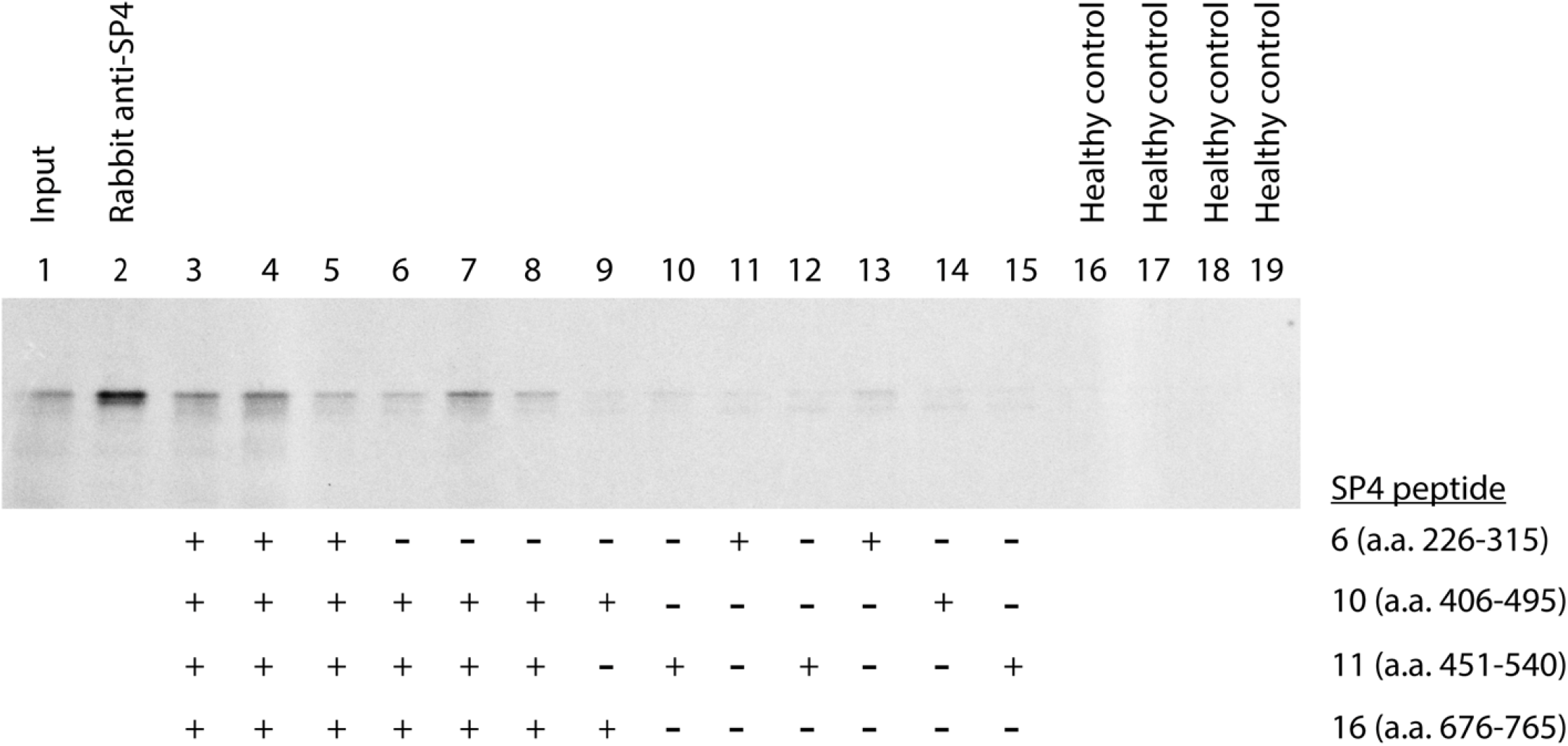
Human serum samples from DM patients immunoprecipitate full-length Sp4 protein. Sera from DM patients that recognized at least one Sp4 peptide by PhIP-Seq were used to immunoprecipitate radiolabeled full-length Sp4 protein. The Sp4 peptides recognized by each serum using PhIP-Seq are indicated below each lane. Those serum samples recognizing 3-4 distinct Sp4 peptides (lanes 3-8) by PhIP-Seq immunoprecipitated full-length Sp4 protein more-efficiently than those serum samples that only recognized 1-2 Sp4 peptides by PhIP-Seq (lanes 9-15). The input Sp4 protein used for immunoprecipitation is shown in lane 1. A commercial rabbit anti-Sp4 autoantibody was used to immunoprecipitate radiolabeled Sp4 in lane 2. Healthy control sera did not immunoprecipitate full-length Sp4 protein (lanes 16-19).

The screening cohort consisted of an additional 321 serum samples from myositis patients seen at the Johns Hopkins Myositis Center. Taken together, the discovery and screening cohorts included sera from 371 myositis patients (255 with DM, 28 with ASyS, 40 with IMNM, 19 with PM, and 29 with IBM). Among these, 29 (7.8%) sera samples were anti-Sp4-positive by ELISA (Figure 2) and all of them had DM. 90% of the anti-SP4-positive DM were TIF1γ-positive (n=26). Out of 39 DM patients with anti-NXP2 autoantibodies and 55 DM patients with anti-Mi2 autoantibodies, 2 (5.1%) and 1 (1.8%) were anti-SP4-positive, respectively. In contrast, no patient with anti-MDA5, anti-SAE, or anti-PMScl autoantibodies also had anti-Sp4 autoantibodies.

**Figure 2.**
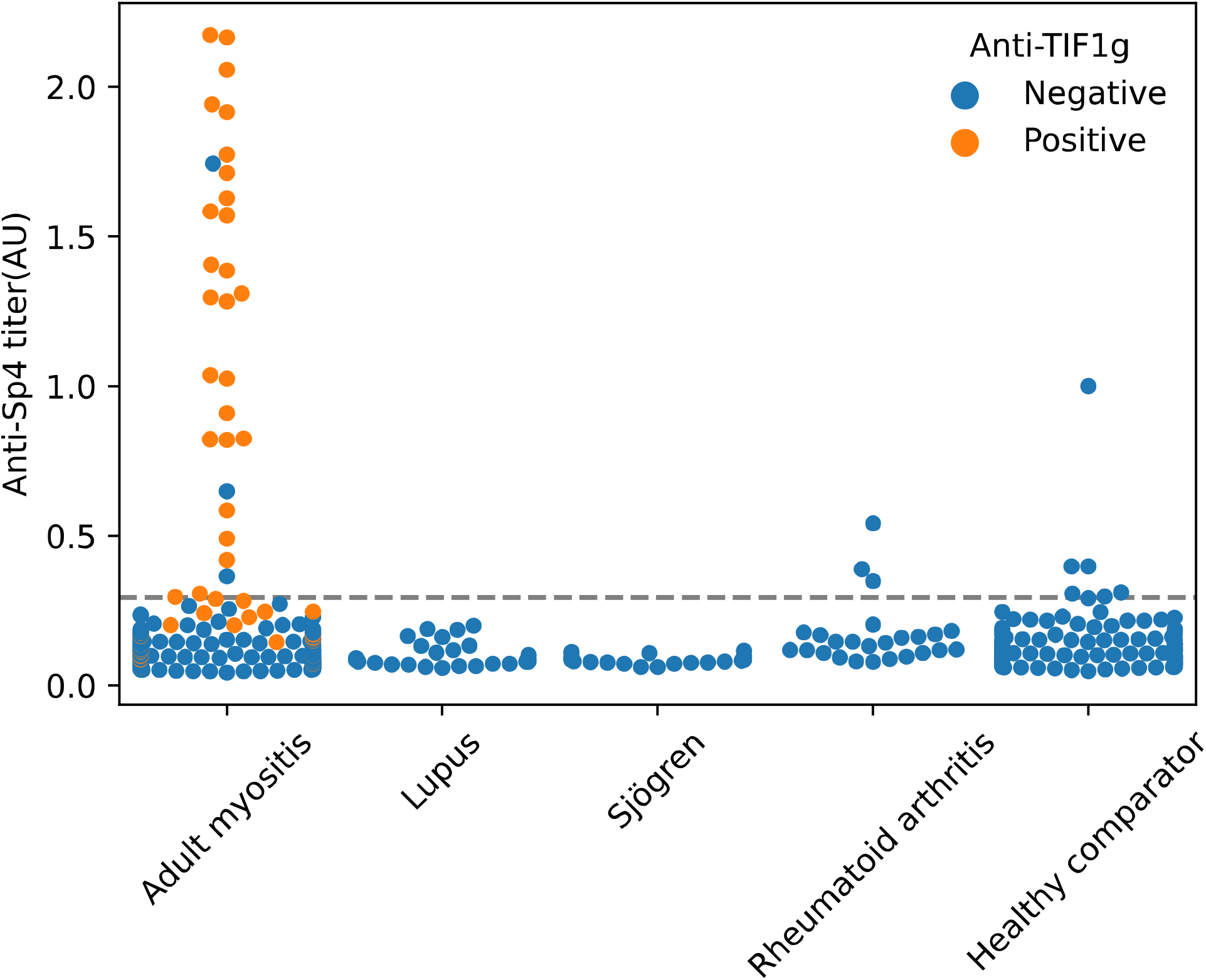
The prevalence of anti-Sp4 autoantibodies in patients with myositis, other rheumatologic conditions, and healthy comparators. Sera from 371 adult myositis patients, 25 lupus patients, 25 Sjogren’s syndrome patients, 25 rheumatoid arthritis patients, and 200 healthy comparators were used in an anti-Sp4 ELISA assay. The dotted line indicates the cut-off used to define anti-Sp4 positive sera.

We also tested for anti-Sp4 autoantibodies in 25 patients with Sjogren’s syndrome, 25 patients with systemic lupus erythematosus, and 25 patients with rheumatoid arthritis. Two (8%) of the rheumatoid arthritis patients were anti-Sp4-positive whereas the rest of the patients with other rheumatologic conditions were negative for these autoantibodies (Figure 2). Taken together, these results indicate that anti-Sp4 autoantibodies can be considered relatively specific for anti-TIF1γ-positive DM.

### The clinical features of TIF1γ-positive DM patients with and without co-existing anti-Sp4 autoantibodies

Most demographic (Table 2) and clinical (Tables 3 and 4) features were similar between TIF1γ-positive DM patients with and without anti-Sp4 autoantibodies. However, patients with anti-Sp4 autoantibodies had measures of muscular strength that were significantly higher than those who were anti-Sp4-negative (Table 4). We also noted that among 26 TIF1γ-positive patients from Johns Hopkins with anti-Sp4 autoantibodies, none had cancer. In contrast, among 35 TIF1γ-positive patients without anti-Sp4 autoantibodies, 5 (14%) had cancer (p=0.04). To confirm that anti-Sp4 autoantibodies are associated with absence of cancer in anti-TIF1γ-positive DM patients, we screened an additional cohort of 23 TIF1γ-positive DM patients from Spain for anti-Sp4 autoantibodies, 13 of whom had cancer. From among 84 total TIF1γ-positive patients, 18 had cancer and none (0%) of these had anti-Sp4 autoantibodies. In contrast, from among 66 patients without cancer, 26 (39%) had anti-Sp4 autoantibodies. Thus, anti-Sp4 autoantibodies are associated with a reduced risk of cancer in anti-TIF1γ-positive DM patients (p=0.001).

**Table 2:** General features of anti-TIF1γ patients with and without anti-Sp4 autoantibodies.

|  | **Anti-Sp4+**<br>**(n=26)** | **Anti-Sp4-**<br>**(n=35)** | **p-value** | **Total**<br>**(n=61)** |
| --- | --- | --- | --- | --- |
| **Female sex** | 81% (21) | 89% (31) | 0.5 | 85% (52) |
| **Race** |  |  |  |  |
| **White** | 85% (22) | 86% (30) | 1.0 | 85% (52) |
| **Black** | 0% (0) | 11% (4) | 0.1 | 7% (4) |
| **Other races** | 15% (4) | 3% (1) | 0.2 | 8% (5) |
| **Age of onset (years)** | 43.0 (13.8) | 50.6 (15.3) | 0.05 | 47.4 (15.1) |
| **Time of follow-up (years)** | 7.1 (4.3) | 4.8 (2.7) | 0.01 | 5.8 (3.7) |
| **Number of visits per participant** | 10.2 (7.7) | 10.9 (6.7) | 0.7 | 10.6 (7.1) |
| **Cancer associated myositis** | 0% (0) | 14% (5) | 0.07 | 8% (5) |
| **Death during follow-up** | 0% (0) | 9% (3) | 0.3 | 5% (3) |
| **Anti-Ro52** | 15% (4) | 17% (6) | 1.0 | 16% (10) |
| **Treatments** |  |  |  |  |
| **Corticosteroids** | 81% (21) | 71% (25) | 0.4 | 75% (46) |
| **Azathioprine** | 27% (7) | 29% (10) | 0.9 | 28% (17) |
| **Methotrexate** | 62% (16) | 54% (19) | 0.6 | 57% (35) |
| **Mycophenolate** | 46% (12) | 46% (16) | 1.0 | 46% (28) |
| **IVIG** | 38% (10) | 57% (20) | 0.1 | 49% (30) |
| **Rituximab** | 12% (3) | 17% (6) | 0.7 | 15% (9) |

*Dichotomous variables were expressed as percentage (count) and continuous variables as mean (SD). Bivariate comparisons of continuous variables were made using Student's t-test while bivariate comparisons of dichotomous variables were made either using chi-squared test or Fisher's exact test, as appropriate.*

**Table 3:** Clinical features of anti-TIF1γ patients with and without anti-Sp4 autoantibodies.

|  | **Anti-Sp4+**<br>**(n=26)** | **Anti-Sp4-**<br>**(n=35)** | **p-value** | **Total**<br>**(n=61)** |
| --- | --- | --- | --- | --- |
| **Muscle involvement** |  |  |  |  |
| **Muscle weakness** | 85% (22) | 80% (28) | 0.7 | 82% (50) |
| **Skin involvement** |  |  |  |  |
| **DM-specific skin involvement** | 100% (26) | 100% (35) | 1 | 100% (61) |
| **Raynaud's phenomenon** | 38% (10) | 17% (6) | 0.06 | 26% (16) |
| **Mechanics hands** | 23% (6) | 26% (9) | 0.8 | 25% (15) |
| **Calcinosis** | 8% (2) | 17% (6) | 0.4 | 13% (8) |
| **Subcutaneous edema** | 8% (2) | 17% (6) | 0.4 | 13% (8) |
| **Lung involvement** |  |  |  |  |
| **Interstitial lung disease** | 4% (1) | 0% (0) | 0.4 | 2% (1) |
| **Esophageal involvement** |  |  |  |  |
| **Gastroesophageal reflux disease** | 27% (7) | 34% (12) | 0.5 | 31% (19) |
| **Dysphagia** | 42% (11) | 54% (19) | 0.4 | 49% (30) |
| **Joint involvement** |  |  |  |  |
| **Arthritis** | 19% (5) | 17% (6) | 1.0 | 18% (11) |
| **Arthralgia** | 58% (15) | 57% (20) | 1.0 | 57% (35) |
| **Systemic involvement** |  |  |  |  |
| **Fever** | 12% (3) | 9% (3) | 1.0 | 10% (6) |

*Chi-squared or Fisher's exact tests were used to compare each one of the clinical groups with the anti-Mi2 patients.*

**Table 4.**
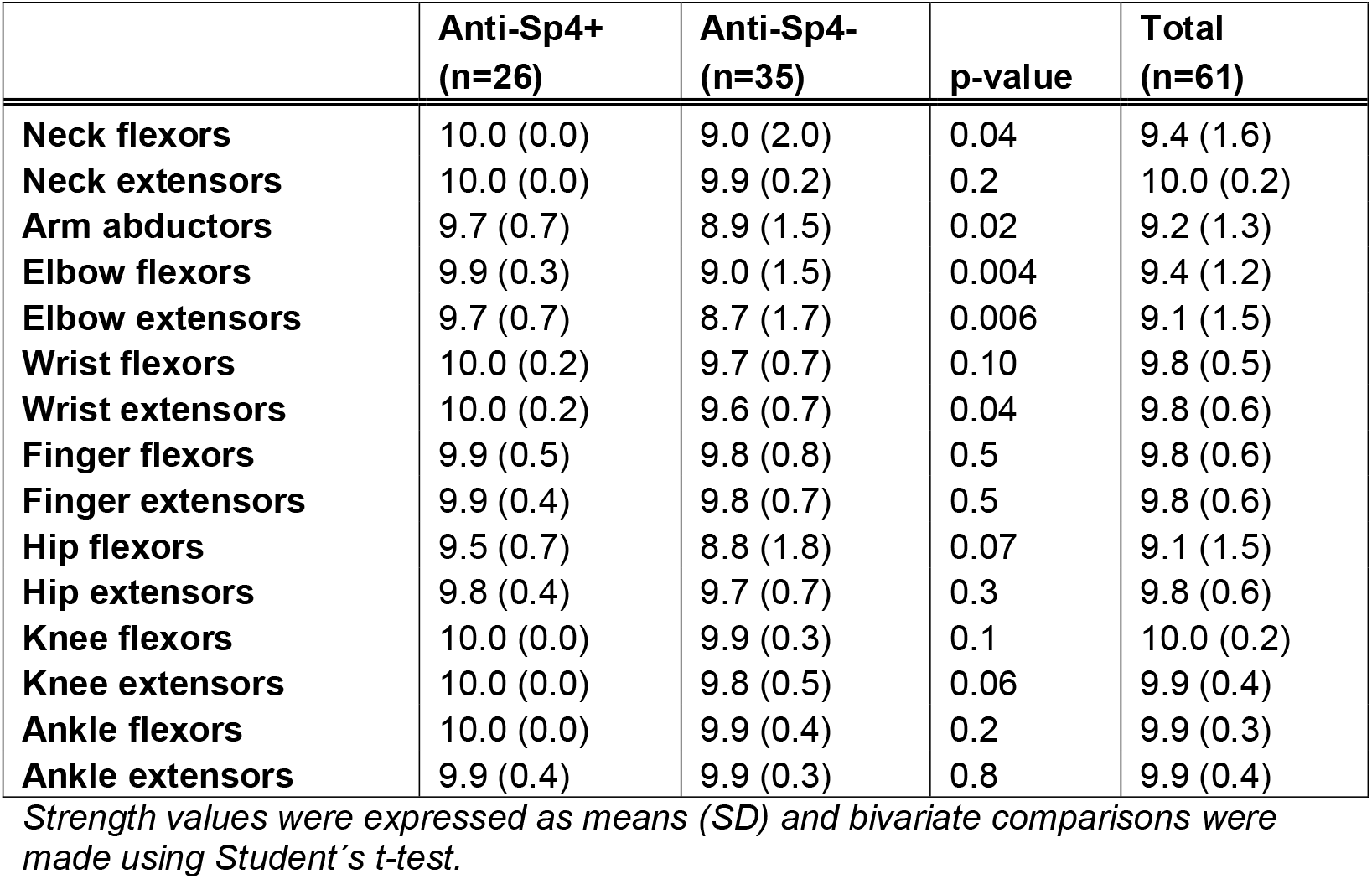
**Weakness pattern at first visit of anti-TIF1γ patients with and without anti-Sp4 autoantibodies**.

|  | **Anti-Sp4+**<br>**(n=26)** | **Anti-Sp4-**<br>**(n=35)** | **p-value** | **Total**<br>**(n=61)** |
| --- | --- | --- | --- | --- |
| **Neck flexors** | 10.0 (0.0) | 9.0 (2.0) | 0.04 | 9.4 (1.6) |
| **Neck extensors** | 10.0 (0.0) | 9.9 (0.2) | 0.2 | 10.0 (0.2) |
| **Arm abductors** | 9.7 (0.7) | 8.9 (1.5) | 0.02 | 9.2 (1.3) |
| **Elbow flexors** | 9.9 (0.3) | 9.0 (1.5) | 0.004 | 9.4 (1.2) |
| **Elbow extensors** | 9.7 (0.7) | 8.7 (1.7) | 0.006 | 9.1 (1.5) |
| **Wrist flexors** | 10.0 (0.2) | 9.7 (0.7) | 0.10 | 9.8 (0.5) |
| **Wrist extensors** | 10.0 (0.2) | 9.6 (0.7) | 0.04 | 9.8 (0.6) |
| **Finger flexors** | 9.9 (0.5) | 9.8 (0.8) | 0.5 | 9.8 (0.6) |
| **Finger extensors** | 9.9 (0.4) | 9.8 (0.7) | 0.5 | 9.8 (0.6) |
| **Hip flexors** | 9.5 (0.7) | 8.8 (1.8) | 0.07 | 9.1 (1.5) |
| **Hip extensors** | 9.8 (0.4) | 9.7 (0.7) | 0.3 | 9.8 (0.6) |
| **Knee flexors** | 10.0 (0.0) | 9.9 (0.3) | 0.1 | 10.0 (0.2) |
| **Knee extensors** | 10.0 (0.0) | 9.8 (0.5) | 0.06 | 9.9 (0.4) |
| **Ankle flexors** | 10.0 (0.0) | 9.9 (0.4) | 0.2 | 9.9 (0.3) |
| **Ankle extensors** | 9.9 (0.4) | 9.9 (0.3) | 0.8 | 9.9 (0.4) |

*Strength values were expressed as means (SD) and bivariate comparisons were made using Student's t-test.*

We next analyzed the evolution of anti-Sp4 autoantibody titers during the disease course by analyzing longitudinally collected serum samples. We found that titers decreased in many patients but did not normalize after treatment of the DM (data not shown). Finally, to determine whether anti-TIF1γ-positive patients without anti-Sp4 autoantibodies at their initial visit to the Johns Hopkins Myositis Center might develop them during the course of the disease, we screened the most recently collected serum samples from 24 TIF1γ-positive and anti-Sp4-negative patients. During an average of 4.7 years (SD 2.3) between the collection of the first and most recent serum samples, only 2 (8.3%) of these became positive for anti-Sp4 autoantibodies.

## DISCUSSION

The association between DM and malignancy has been appreciated for over a century^20, 21^. In more recent years, the presence of autoantibodies against TIF1γ has been recognized to be associated with an especially high risk of malignancy in DM patients. Indeed, one recent meta-analysis including a total of 312 patients showed that 80% of patients with cancer-associated DM had anti-TIF1γ autoantibodies whereas only 10% without cancer were anti-TIF1γ-positive^2^. Overall, the presence of anti-TIF1γ autoantibodies had a positive predictive value of 58% and a negative predictive value of 93% for diagnosing cancer-associated DM^2^. Interestingly, it now appears that anti-TIF1γ autoantibodies of different IgG subclasses have variable cancer risks. Specifically, a recent study revealed that the IgG2 subclass is closely associated with malignancy and 100% of patients with high titer anti-TIF1γ IgG2 autoantibodies developed cancer within 24 months of the DM diagnosis^22^.

Although the nature of the relationship between malignancy and autoantibodies recognizing TIF1γ is incompletely understood, several lines of evidence raise the possibility that anti-TIF1γ autoimmunity may be induced by the tumor. First, tumors from anti-TIF1γ-positive patients have an increased prevalence of somatic mutations and/or loss of heterozygosity in their TIF1 genes when compared with tumors from TIF1γ-negative patients^23^. The loss of heterozygosity is consistent with immunoediting due to an immune response directed against tumor cells with mutations in TIF1γ. Second, TIF1γ protein levels appear to be increased in tumors from anti-TIF1γ-positive DM patients compared to tumors from non-myositis patients^23^, suggesting that that increased expression of the protein in the tumor may play a role in breaking immune tolerance. Third, in anti-TIF1γ-positive DM patients who have undergone successful treatment of their malignancy, the autoantibodies can become undetectable, and their DM may enter remission^24^.

Despite the strong link between anti-TIF1γ autoantibodies and cancer, a malignancy never emerges in approximately 40% of DM patients with this autoantibody^2^. Recently, autoantibodies against cell division cycle and apoptosis regulator 1 (CCAR1) protein were found to be associated with decreased cancer prevalence among anti-TIF1γ-positive DM patients; they were present in 36% of those without cancer and in 22% of those with cancer^25^. In the current study, we discovered novel autoantibodies against Sp4 protein that appear to have a markedly stronger association with the absence of cancer in anti-TIF1γ-positive DM patients. Among the TIF1γ-positive DM patients in the current study, anti-Sp4 autoantibodies were present in 39% without cancer and in 0% of those with cancer. Sp4 is a transcription factor that is not only important for early development but, like TIF1γ, is also expressed at high levels in various tumors^26^. Future experiments will determine if TIF1γ and Sp4 are components of a molecular complex or otherwise interact biochemically. If so, it may be that Sp4 antibodies serve as a surrogate for epitope spreading during a more vigorous anti-tumor immune response.

Interestingly, patients with both anti-TIF1γ and anti-Sp4 autoantibodies do not appear to have more severe or advanced DM compared with patients harboring anti-TIF1γ autoantibodies in isolation. Indeed, patients with both autoantibodies exhibited significantly better arm strength than those with just TIF1γ autoantibodies. Of note, an analogous phenomenon has been observed in systemic sclerosis, where patients with autoantibodies against RNA polymerase III have an increased risk of cancer, but those with co-existing anti-RNA polymerase I autoantibodies do not^27^. And similar to what was observed in DM, systemic sclerosis patients with both autoantibodies had no evidence of more severe autoimmune disease than those with anti-RNA polymerase III autoantibodies alone.

Future prospective studies will be required to determine whether DM patients with both anti-TIF1γ and anti-Sp4 autoantibodies require less aggressive malignancy screening. Moreover, additional studies will be required to demonstrate whether or not the anti-Sp4 immune response directly damages tumor cells and, if so, whether this effect might be harnessed to therapeutic advantage in non-myositis patients with cancer.

## Data Availability

All data relevant to the study are either included in the article or will be shared upon request.

## Acknowledgments

This work was supported, in part, by the Intramural Research Program of the National Institutes of Arthritis and Musculoskeletal and Skin Diseases of the National Institutes of Health. The Johns Hopkins Rheumatic Diseases Research Core Center, where some of the autoantibodies were assayed, is supported by NIH P30-AR070254.

## Notes

### Competing Interest Statement

The authors have declared no competing interest.

### Author Declarations

IRB of Johns Hopkins gave ethical approval for this work. IRB of the National Institutes of Health gave ethical approval for this work. Ethics committee of the Vall dHebron and Clinic Hospitals gave ethical approval for this work.

## REFERENCES

1. Selva-O’Callaghan A, Pinal-Fernandez I, Trallero-Araguas E, Milisenda JC, Grau-Junyent JM, Mammen AL. Classification and management of adult inflammatory myopathies. Lancet Neurol 2018;17:816–828.

2. Trallero-Araguas E, Rodrigo-Pendas JA, Selva-O’Callaghan A, et al. Usefulness of anti-p155 autoantibody for diagnosing cancer-associated dermatomyositis: a systematic review and meta-analysis. Arthritis and Rheumatism 2012;64:523–532.

3. Pinal-Fernandez I, Mecoli CA, Casal-Dominguez M, et al. More prominent muscle involvement in patients with dermatomyositis with anti-Mi2 autoantibodies. Neurology 2019.

4. Kochi Y, Kamatani Y, Kondo Y, et al. Splicing variant of WDFY4 augments MDA5 signalling and the risk of clinically amyopathic dermatomyositis. Ann Rheum Dis 2018;77:602–611.

5. Larman HB, Laserson U, Querol L, et al. PhIP-Seq characterization of autoantibodies from patients with multiple sclerosis, type 1 diabetes and rheumatoid arthritis. J Autoimmun 2013;43:1–9.

6. Larman HB, Zhao Z, Laserson U, et al. Autoantigen discovery with a synthetic human peptidome. Nat Biotechnol 2011;29:535–541.

7. Bohan A, Peter JB. Polymyositis and dermatomyositis (first of two parts). The New England journal of medicine 1975;292:344–347.

8. Bohan A, Peter JB. Polymyositis and dermatomyositis (second of two parts). The New England journal of medicine 1975;292:403–407.

9. Mohan D, Wansley DL, Sie BM, et al. PhIP-Seq characterization of serum antibodies using oligonucleotide-encoded peptidomes. Nat Protoc 2018;13:1958–1978.

10. Mammen AL, Chung T, Christopher-Stine L, et al. Autoantibodies against 3-hydroxy-3-methylglutaryl-coenzyme A reductase in patients with statin-associated autoimmune myopathy. Arthritis Rheum 2011;63:713–721.

11. Amici DR, Pinal-Fernandez I, Mazala DA, et al. Calcium dysregulation, functional calpainopathy, and endoplasmic reticulum stress in sporadic inclusion body myositis. Acta Neuropathol Commun 2017;5:24.

12. Targoff IN, Mamyrova G, Trieu EP, et al. A novel autoantibody to a 155-kd protein is associated with dermatomyositis. Arthritis Rheum 2006;54:3682–3689.

13. Rider LG, Werth VP, Huber AM, et al. Measures of adult and juvenile dermatomyositis, polymyositis, and inclusion body myositis: Physician and Patient/Parent Global Activity, Manual Muscle Testing (MMT), Health Assessment Questionnaire (HAQ)/Childhood Health Assessment Questionnaire (C-HAQ), Childhood Myositis Assessment Scale (CMAS), Myositis Disease Activity Assessment Tool (MDAAT), Disease Activity Score (DAS), Short Form 36 (SF-36), Child Health Questionnaire (CHQ), physician global damage, Myositis Damage Index (MDI), Quantitative Muscle Testing (QMT), Myositis Functional Index-2 (FI-2), Myositis Activities Profile (MAP), Inclusion Body Myositis Functional Rating Scale (IBMFRS), Cutaneous Dermatomyositis Disease Area and Severity Index (CDASI), Cutaneous Assessment Tool (CAT), Dermatomyositis Skin Severity Index (DSSI), Skindex, and Dermatology Life Quality Index (DLQI). Arthritis Care Res (Hoboken) 2011;63 Suppl 11:S118–157.

14. Travis WD, Costabel U, Hansell DM, et al. An official American Thoracic Society/European Respiratory Society statement: Update of the international multidisciplinary classification of the idiopathic interstitial pneumonias. Am J Respir Crit Care Med 2013;188:733–748.

15. Xu GJ, Shah AA, Li MZ, et al. Systematic autoantigen analysis identifies a distinct subtype of scleroderma with coincident cancer. Proc Natl Acad Sci U S A 2016;113:E7526–E7534.

16. Mohan D, Wansley DL, Sie BM, et al. Publisher Correction: PhIP-Seq characterization of serum antibodies using oligonucleotide-encoded peptidomes. Nat Protoc 2019;14:2596.

17. Mammen AL, Chung T, Christopher-Stine L, et al. Autoantibodies against 3-hydroxy-3-methylglutaryl-coenzyme A reductase in patients with statin-associated autoimmune myopathy. Arthritis and Rheumatism 2011;63:713–721.

18. Larman HB, Salajegheh M, Nazareno R, et al. Cytosolic 5’-nucleotidase 1A autoimmunity in sporadic inclusion body myositis. Annals of Neurology 2013;73:408–418.

19. Espinosa-Ortega F, Holmqvist M, Alexanderson H, et al. Comparison of autoantibody specificities tested by a line blot assay and immunoprecipitation-based algorithm in patients with idiopathic inflammatory myopathies. Ann Rheum Dis 2019;78:858–860.

20. Stertz G. Polymyositis. Berl Klin Wochenschr 1916;53:489.

21. Tiniakou E, Mammen AL. Idiopathic Inflammatory Myopathies and Malignancy: a Comprehensive Review. Clin Rev Allergy Immunol 2017;52:20–33.

22. Aussy A, Freret M, Gallay L, et al. The IgG2 isotype of anti-transcription intermediary factor 1-gamma autoantibodies is a biomarker of mortality in adult dermatomyositis. Arthritis Rheumatol 2019.

23. Pinal-Fernandez I, Ferrer-Fabregas B, Trallero-Araguas E, et al. Tumour TIF1 mutations and loss of heterozygosity related to cancer-associated myositis. Rheumatology (Oxford) 2018;57:388–396.

24. Dani L, Holmqvist M, Martinez MA, et al. Anti-transcriptional intermediary factor 1 gamma antibodies in cancer-associated myositis: a longitudinal study. Clin Exp Rheumatol 2019.

25. Fiorentino DF, Mecoli CA, Rosen MC, et al. Immune responses to CCAR1 and other dermatomyositis autoantigens are associated with attenuated cancer emergence. J Clin Invest 2022;132.

26. Safe S, Abbruzzese J, Abdelrahim M, Hedrick E. Specificity Protein Transcription Factors and Cancer: Opportunities for Drug Development. Cancer Prev Res (Phila) 2018;11:371–382.

27. Shah AA, Laiho M, Rosen A, Casciola-Rosen L. Protective Effect Against Cancer of Antibodies to the Large Subunits of Both RNA Polymerases I and III in Scleroderma. Arthritis Rheumatol 2019;71:1571–1579.

